# Severity of COVID-19 reinfection and associated risk factors: findings of a cross-sectional study in Bangladesh

**DOI:** 10.1101/2021.12.26.21268408

**Authors:** Md. Ziaul Islam, Baizid Khoorshid Riaz, Shah Ali Akbar Ashrafi, Sharmin Farjana, Syeda Sumaiya Efa, Mohammad Adnan Khan

## Abstract

**Background:** COVID-19 reinfected patients suffer from diverse health consequences. Information on the severity of COVID-19 reinfection is scarce. The current study aimed to determine the proportion of COVID-19 reinfection and risk factors associated with its severity.

**Methods:** This cross-sectional study targeted all COVID-19 patients reported in May 2021 at the Health Information Unit (HIU) of the Directorate General of Health Services (DGHS) of Bangladesh. We identified 473 (1.14%) reinfected patients out of 41408 diagnosed cases by reviewing their medical records. Considering the selection criteria and informed consent, we enrolled 404 reinfected patients. Data were collected through telephone interviews and reviewing medical records using a semi-structured questionnaire and a checklist.

**Results:** The majority of the reinfected patients were urban residents (98.0%). Around 13.0% of reinfected patients had <90% oxygen saturation, and 64.0% had an interval of 3-6 months between two attacks. The severity of reinfection included asymptomatic (12.9%), mild (8.9%), moderate (66.3%), and severe (11.9%) forms of infection. An interval of 3-6 months between two attacks had less chance of having mild (AOR=0.031, ρ=0.000), moderate (AOR=0.132, ρ=0.017), and severe (AOR=0.059, ρ=0.002) infections. Patients who maintained physical distance had less chance of moderate-intensity reinfection (AOR=0.137, ρ=0.013), while the vaccinated patients had a higher chance of moderate (AOR=16.127, ρ=0.001) and severe (AOR=3.894, ρ=0.047) intensity reinfection.

**Conclusion:** To avert COVID-19 reinfection and its severity, patients should be vigilant about preventive practices even after recovery. The study suggests vibrant interventions aligned with exposure, physical distancing, vaccination, and comorbidities for mitigating reinfection.

## Introduction

In over 200 nations, the Coronavirus Disease 2019 (COVID-19) pandemic has infected over 23 million patients, resulting in over 0.8 million deaths. The pandemic has impacted negatively on the healthcare system and put a stop to socioeconomic activities [1]. COVID-19 was thought to be a disease caused by a stable virus that could provide immunity; similar to most respiratory viruses (with the notable exception of rhinoviruses) who provide immunity for a year or more. But it gradually became obvious that naturally acquired immunity of COVID-19 would not always provide security in the months following the initial infection. This may be due to a lack of effective natural immunity following infection or the presence of variants on major epitopes that could potentially contribute to infection resistance [2].

With the growing number of COVID -19 cases around the world, the main concern, apart from the vaccine, is several episodes of coronavirus infection in a single person, also known as COVID-19 reinfection [3]. Many countries around the world have confirmed reinfections which raised questions on the vaccine’s prospects and ability to protect the public from the disease [4].

SARS□CoV□2 reactivation or reinfection is a persistent and vexing problem and also a major public health concern in terms of global morbidity and mortality. There is a possibility that recurrence of SARS□CoV□2 infection may occur from false-negative RT□PCR results which creates the need for a longer observation period for recovered COVID□19 patients. The possibility of discharged patients suffering reactivation or being re□infected with another SARS□CoV□2 strain also cannot be excluded [5].

Depending on the rate and type of immune response, severity and rate of reinfection may vary from society to society [6]. A recently study showed that most people infected with COVID-19 showed antibody response between 10 and 14 days after infection whereas in some mild cases, detection of antibodies requires a long time after symptoms, and in few cases, antibodies are not detected at all. There is a lack of information regarding the longevity of the antibody response to SARS-CoV-2, but it is known that antibodies to other human coronaviruses wane over time, and reinfection occurs. Thus, reinfection of previously infected COVID-19 cases is a real possibility that should be considered in the models of the post-pandemic era [7].

Based on the findings of some studies regarding the evidence of prolonged viral shedding up to 82 days, the Centers for Disease Control and Prevention (CDC) has considered 90 days between two positive SARS-CoV-2 RNA tests along with genomic evidence of reinfection as an investigative criterion to understand the phenomenon of reinfection [8]. According to European CDC (2020), reinfection is defined as laboratory confirmation of two infections by two different strains (minimum distance to be determined or supported by phylogenetic and epidemiological data) with timely separated illness/infection episodes [9].

No concrete evidence is available regarding the factors associated with COVID-19 reinfection. Some studies identified that fatigue, positive IgM, positive IgG, lower platelet count, etc., are associated with an increased risk of recurrent infection [10]. Moreover, some recent studies did not support the possibility of COVID-19 reinfection after 70 days following the initial infection and the evidence for COVID-19 recurrence due to viral relapse [11]. A very little information is available regarding the factors which influence the severity of COVID-19 reinfection.

Although several cases of COVID-19 reinfection have been reported in Bangladesh but concrete data on proportion, severity, and associated risk factors are not available. The present study aimed to determine the occurrence of COVID-19 reinfection and the risk factors associated with severity.

## Methods

### Study setting, design, and subjects

This countrywide cross-sectional study was conducted at the Health Information Unit (HIU) of the Directorate General Health Services (DGHS), Bangladesh, from March to August 2021. The target population was reinfected COVID-19 patients diagnosed by real time reverse transcription polymerase chain reaction (RT-PCR) assay who reported to the HIU of the DGHS in May 2021. A total of 41408 COVID-19 patients reported to the HIU of the DGHS in May 2021. We reviewed the medical records of all the COVID-19 patients to find out the reinfected patients irrespective of their age, sex, and residence. We approached all the reinfected patients for participating in the study. The participants who did not respond to a phone call on three (03) separate occasions; who were unwilling to participate and severely ill were excluded from the study.

### Sample size and sampling

With the administrative permission of HIU, DGHS, medical records of all the COVID-19 patients (41408 who were reported in May 2021) were reviewed. Among all the diagnosed COVID-19 patients, 473 patients were identified as the reinfected cases. We reviewed the present and previous history of illness, clinical findings, and socio-demographic characteristics including address and contact number of the reinfected patients. Relevant medical records were collected from the HIU, DGHS. Due to failure of mobile contact or unwillingness or incomplete response, a total of 404 reinfected COVID-19 patients were enrolled in the study. All the patients were selected purposively and enrolled in the study voluntarily.

### Data collection instrument and methods

Data were collected through telephone interviews and reviewing medical records using a semi-structured questionnaire and a checklist respectively. The data collection instruments were pretested, and accordingly, necessary corrections were done for its’ finalization. The semi-structured questionnaire was used to collect information on socio-demographic characteristics, clinical features related to COVID-19 reinfection, the severity of COVID-19 reinfection, comorbidities, and information related to social exposure, preventive practices, and vaccination of COVID-19 reinfected patients. The checklist was used to collect information by reviewing the medical records of patients. Data were collected by the skilled data enumerators, who were trained for five days on the research procedure, data collection instruments and techniques, and quality control of data. Each telephone interview session was recorded by a digital recorder to preserve the consents of the participants and to ensure the validity of the data.

### Measurements

We considered the operational definition of ‘reinfection’ as ‘clinical recurrence of symptoms compatible with COVID-19, accompanied by positive RT-PCR test (cycle threshold value, Ct <35), more than 90 days after the onset of the primary infection, supported by close-contact exposure or outbreak settings, and no evidence of another cause of infection [12].

The severity of COVID-19 reinfection was categorized into asymptomatic, mild, moderate, severe, and critical illness according to the clinical presentation of the patients (NIH, 2021). The asymptomatic patients included those individuals who were test-positive for SARS-CoV-2 using a virologic test but who had no symptoms that are consistent with COVID-19 [13].

According to the national guideline of Bangladesh on clinical management of COVID-19 patients the severity of COVID-19 was categorized into mild, moderate, severe, and critical [14]. The mild illness included those individuals whose clinical symptoms were mild, and there was no evidence of pneumonia [14]. On the other hand, the moderate illness included those individuals who showed clinical signs of pneumonia (fever, cough, dyspnea, fast breathing) but had no sign of severe pneumonia, peripheral capillary *oxygen saturation (*SpO2) ≥ 90% on room air [14].

The severe illness comprised of the individuals who had clinical signs of pneumonia (fever, cough, dyspnea, fast breathing) plus one of the following: severe respiratory distress, respiratory rate > 30 breaths/min or SpO2 < 90% on room air [14]. On the contrary, the critical illness (Cases requiring ICU care) included the severe COVID-19 cases meeting any of the following criteria: respiratory failure and requiring mechanical ventilation; sepsis; septic shock; ARDS; any organ failure that requires ICU care [14].

Patients were regarded as vaccinated who had taken at least one dose of vaccine. Having comorbidity comprised of the patients who had at least one of the comorbidities including ischemic heart disease, hypertension, diabetes mellitus, chronic obstructive pulmonary disease, chronic kidney disease, chronic liver disease, asthma, malignant disease, and hypothyroidism. Those patients were considered as ‘practicing preventive measures’ who maintained all the preventive measures including using a mask during outdoor activities and attending to people, washing hands and using hand sanitizer, and maintaining physical distance regularly.

### Statistical analysis

All responses were checked for their completeness, correctness to exclude missing or inconsistent data. All collected data were compiled together using the Statistical Package for the Social Science (SPSS) software (Version 25.0, IBM Statistical Product and Service Solutions, Armonk, NY, USA). Data analysis included descriptive statistics (frequency distribution, percentage, mean and standard deviation) and inferential statistics (chi-square test, Fisher’s Exact test, t-test, and multi-nominal logistics regression). Necessary tabulations were drawn for summarizing and smooth visual presentation of data.

The normality of the variables was tested with the Shapiro Wilk test/Kolmogorov Smirnov tests of Normality. Continuous data were written in the form of mean and standard deviation. Categorical variables were reported as frequencies and percentages. Group comparisons of the variables were made and an association was tested with the Chi-square test or Fisher’s exact test as appropriate. Finally, the strength of association among the variables was analysed by multi-nominal logistic regression. A ρ value < 0.05 was considered significant. All the statistical tests were two-sided and were performed at 95% confidence interval (CI) with a significance level of α = 0.05.

### Ethical Issues

The study was conducted considering all kinds of ethical issues in different stages of the study. The ethical approval was obtained from the Institutional Review Board (IRB) of the National Institute of Preventive and Social Medicine (NIPSOM), Dhaka, Bangladesh. Keeping compliance with Helsinki Declaration for medical research involving human subjects, we obtained informed verbal consent from each participant and recorded it in a digital recorder with the permission of the respective participant. Each participant was enrolled voluntarily and explained about the study design, purpose, procedure, risk, and benefits before the interview. We maintained privacy, anonymity, and confidentiality of data strictly. The participants had the right to withdraw their consent at any stage of the study.

## Results

The proportion of COVID-19 reinfection was 1.14% (473 out of 41,408). Among 473 reinfected COVID-19 patients, 85.4% participated in the study, 6.6% didn’t attend the telephone calls, 4.4% were unwilling to participate, and 3.6% had an incomplete interview (Fig. 1).

**Fig 1.**
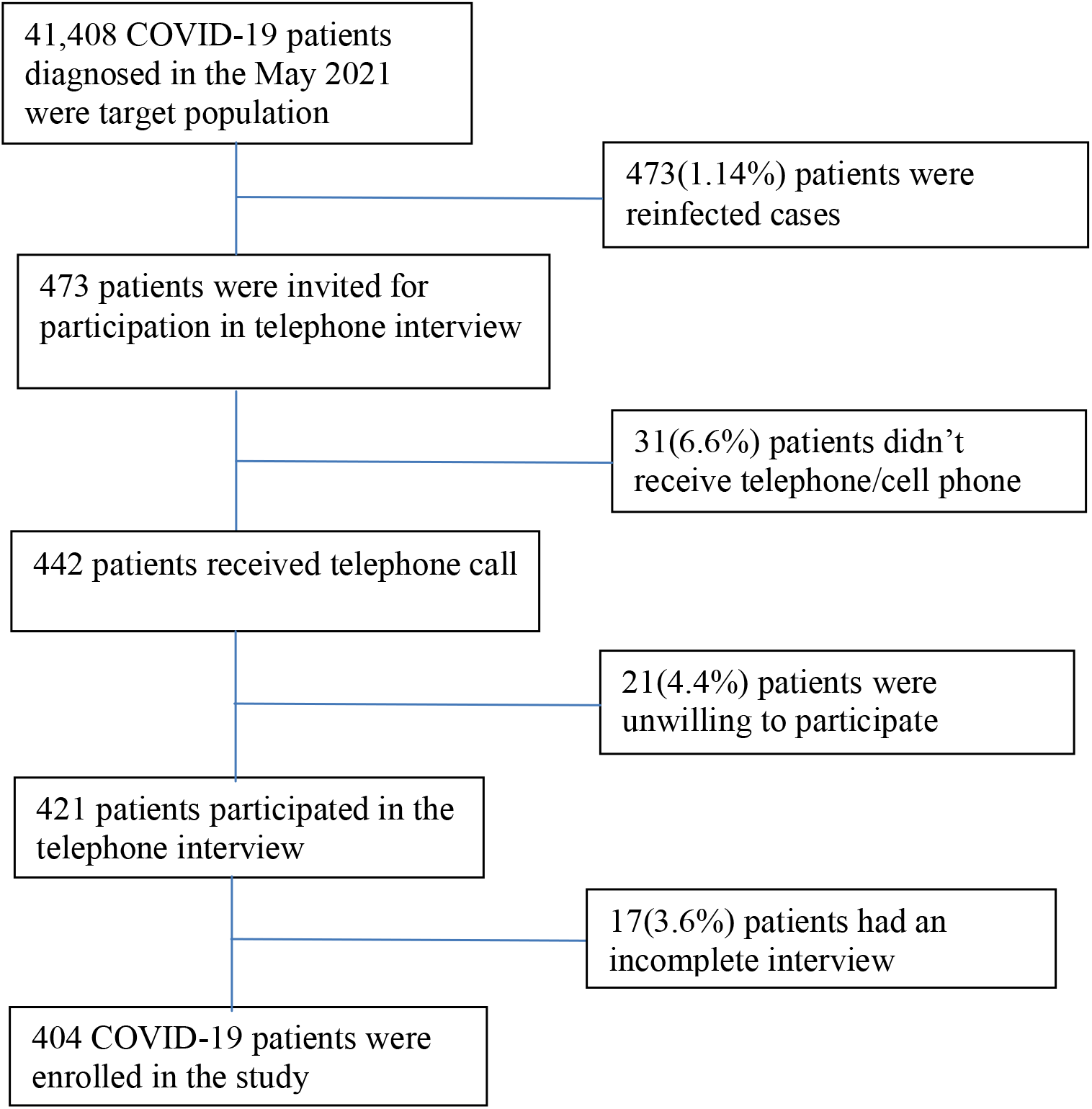
Flowchart of the study participants (COVID-19 reinfected patients)

Among the 404 re-infected COVID-19 patients, the majority (51.0%) were in the age group of 30-39 years. More than half (52.0%) of the patients were male, and the majority (79.7%) were married. Of all, 37.1% were healthcare workers, and 29.2% were in other services. Most (98.0%) of the patients were urban residents, and a majority (39.6%) of the patients had monthly family income within the range of US$578 - 1165 (Table-1).

**Table 1.**
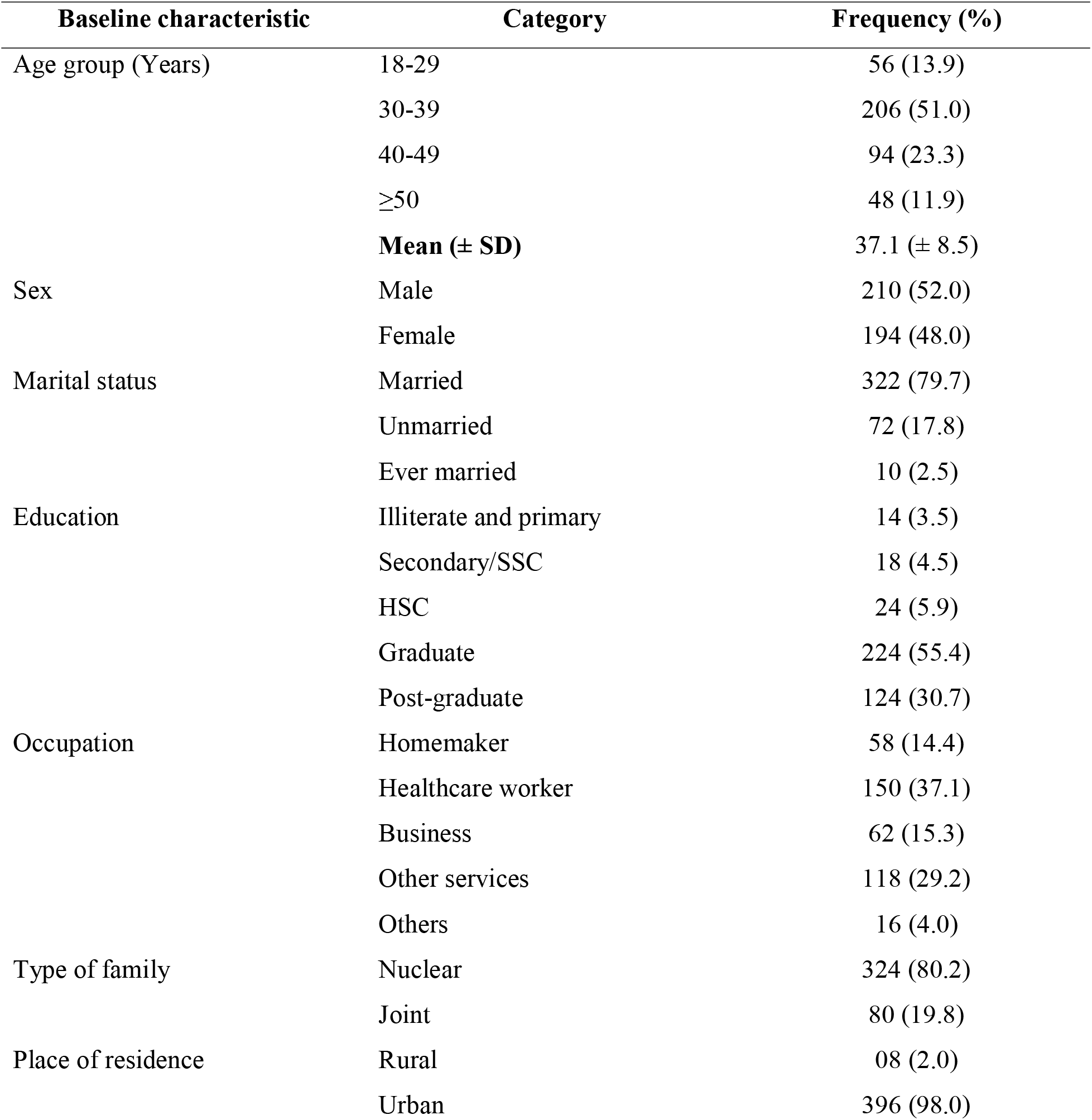
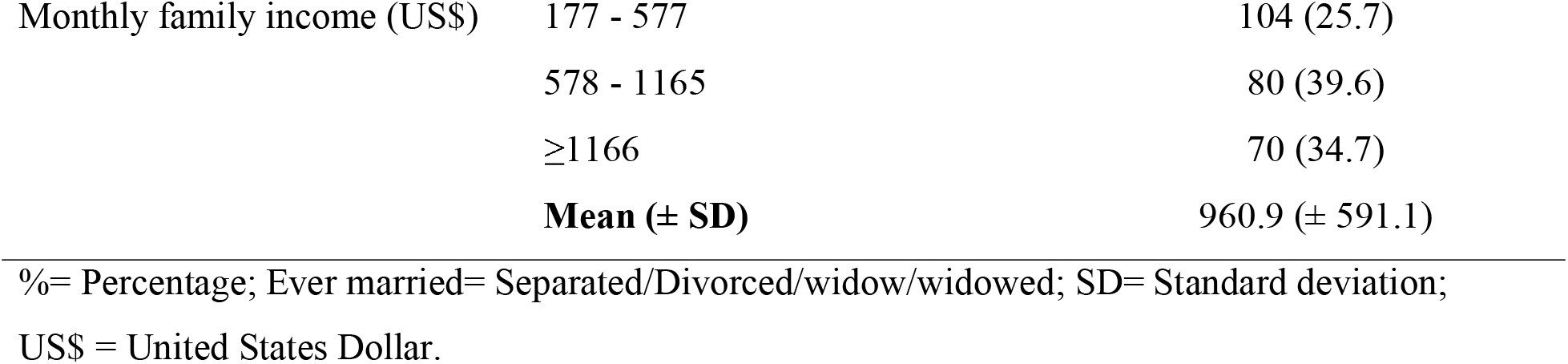
Baseline characteristics of the re-infected COVID-19 patients (n=404)

| Baseline characteristic | Category | Frequency (%) |
| --- | --- | --- |
| Age group (Years) | 18-29 | 56 (13.9) |
|  | 30-39 | 206 (51.0) |
|  | 40-49 | 94 (23.3) |
|  | ≥50 | 48 (11.9) |
|  | **Mean (± SD)** | 37.1 (± 8.5) |
| Sex | Male | 210 (52.0) |
|  | Female | 194 (48.0) |
| Marital status | Married | 322 (79.7) |
|  | Unmarried | 72 (17.8) |
|  | Ever married | 10 (2.5) |
| Education | Illiterate and primary | 14 (3.5) |
|  | Secondary/SSC | 18 (4.5) |
|  | HSC | 24 (5.9) |
|  | Graduate | 224 (55.4) |
|  | Post-graduate | 124 (30.7) |
| Occupation | Homemaker | 58 (14.4) |
|  | Healthcare worker | 150 (37.1) |
|  | Business | 62 (15.3) |
|  | Other services | 118 (29.2) |
|  | Others | 16 (4.0) |
| Type of family | Nuclear | 324 (80.2) |
|  | Joint | 80 (19.8) |
| Place of residence | Rural | 08 (2.0) |
|  | Urban | 396 (98.0) |
| Monthly family income (US\$) | 177 - 577 | 104 (25.7) |
|  | 578 - 1165 | 80 (39.6) |
|  | ≥1166 | 70 (34.7) |
|  | **Mean (± SD)** | **960.9 (± 591.1)** |

%= Percentage; Ever married= Separated/Divorced/widow/widowed; SD= Standard deviation; US\$ = United States Dollar.

**Table 2.**
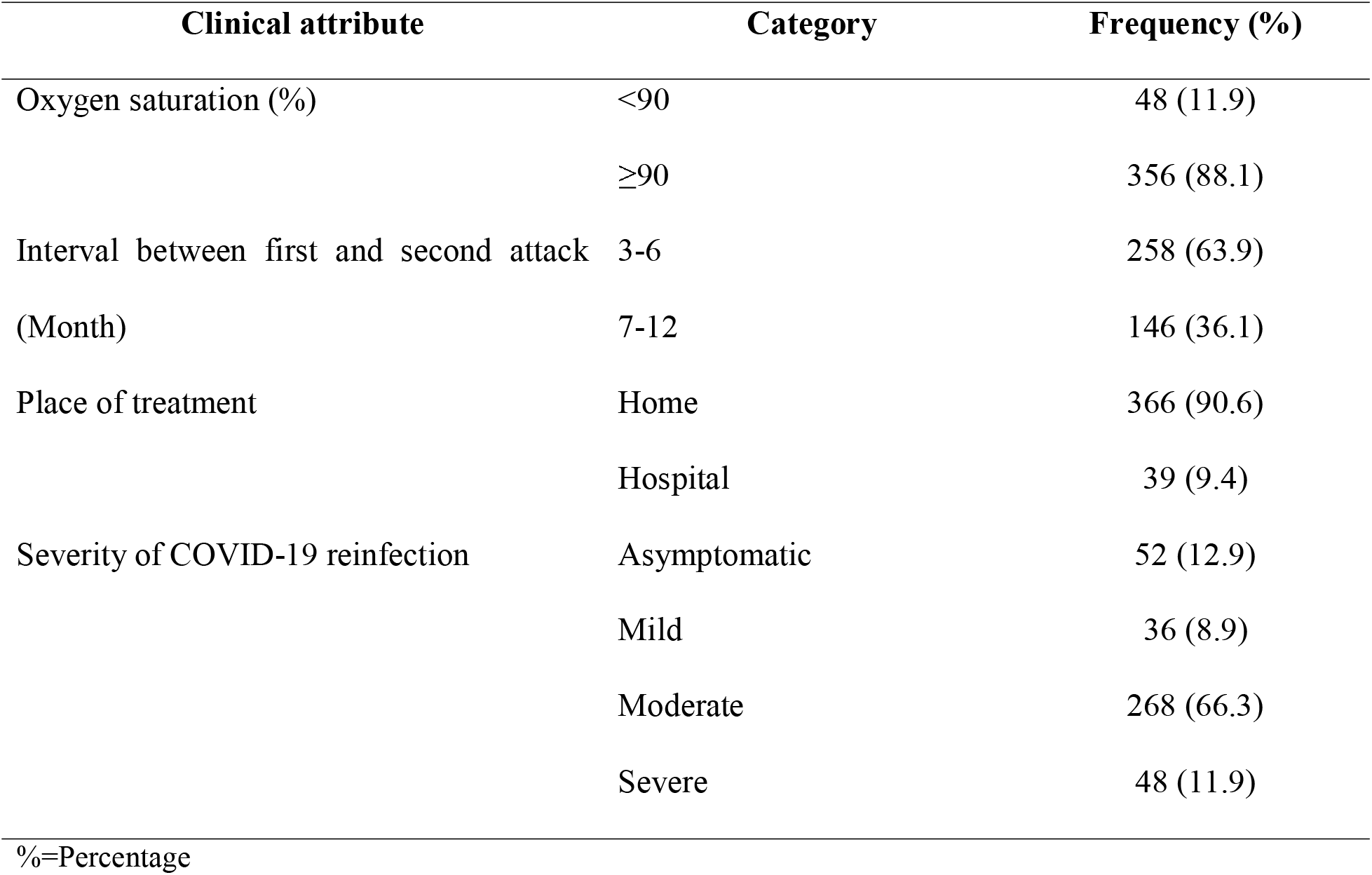
Distribution of COVID-19 reinfected patients by clinical attributes (n=404)

| Clinical attribute | Category | Frequency (%) |
| --- | --- | --- |
| Oxygen saturation (%) | <90 | 48 (11.9) |
|  | ≥90 | 356 (88.1) |
| Interval between first and second attack (Month) | 3-6 | 258 (63.9) |
|  | 7-12 | 146 (36.1) |
| Place of treatment | Home | 366 (90.6) |
|  | Hospital | 39 (9.4) |
| Severity of COVID-19 reinfection | Asymptomatic | 52 (12.9) |
|  | Mild | 36 (8.9) |
|  | Moderate | 268 (66.3) |
|  | Severe | 48 (11.9) |

%=Percentage

**Table 3.** Different categories of risk factors associated with COVID-19 reinfection (n=404) Category Associated factors f (%)

| Category | Associated factors | f (%) |
| --- | --- | --- |
| Exposure related | Performed outside activities | 382 (94.6) |
|  | Attended social gathering | 254 (62.9) |
|  | Used public transport | 154 (38.1) |
| Preventive practice related | Didn't maintain physical distance | 366 (90.6) |
|  | Didn't use mask | 10 (2.5) |
|  | Didn't sanitize hand | 06 (1.5) |
| Having comorbidity | Yes | 116 (28.7) |
|  | No | 288 (71.3) |
| Type of comorbidity | CVD | 12 (3.0) |
|  | HTN | 94 (23.3) |
|  | DM | 84 (20.8) |
|  | COPD | 06 (1.5) |
|  | Hypothyroidism | 16 (4.0) |
| Smoking history | Yes | 108 (26.7) |
|  | No | 296 (73.3) |
| COVID-19 Vaccinated | Yes | 159 (39.1) |
|  | No | 246 (60.9) |

%; Percentage; CVD: Cardiovascular disease; HTN: Hypertension; DM: Diabetes Mellitus; COPD: Chronic obstructive Pulmonary Disease

The majority (88.1%) had ≥90% while 12.9% had <90% oxygen saturation. Around two-thirds of the patients (63.9%) had an interval of 3-6 months between first and second attack, and most (90.6%) of the patients were treated at home. In respect of severity of COVID-19 reinfection, 12.9% were asymptomatic, followed by 66.3% had moderate, 11.9% had severe, and only 8.9% had a mild infection (Table-1).

Regarding the history of exposure, most (94.6%) of the reinfected patients attended outside activities, 62.9% attended the social gathering, and 38.1% used public transports. Regarding preventive practices, most (90.6%) of the patients didn’t maintain physical distance, 2.5% didn’t use a mask, and 1.5% didn’t sanitize their hands. The study found that 28.7% of patients had comorbidities. Among all, 23.3% had hypertension (HTN), 20.8% had diabetes mellitus (DM), and 26.7% had the habit of smoking. Around two-thirds of (60.9%) were not vaccinated against COVID-19 (Table-1).

The severity of COVID-19 reinfection was significantly (ρ = 0.005) associated with age; 74.5% of the age group 40-49 years had moderate while 14.6% of the age group 30-39 years had a severe infection. The severity of reinfection was significantly (ρ<0.001) associated with interval between first and second attack; 68.2% of the patients with an interval of 3-6 months had moderate and 16.4% of the patients with an interval of 7-12 months had a severe infection. The severity of reinfection was significantly associated with maintaining physical distance (ρ<0.001). It moderate infection was significantly higher among the patients who didn’t maintain physical distance (69.9%) than those who maintained physical distance (31.6%). On the contrary, severe infection was significantly higher in the patients who maintained (47.4%) than in the patients who didn’t maintain (8.2%) physical distance (Table 4).

**Table 4.**
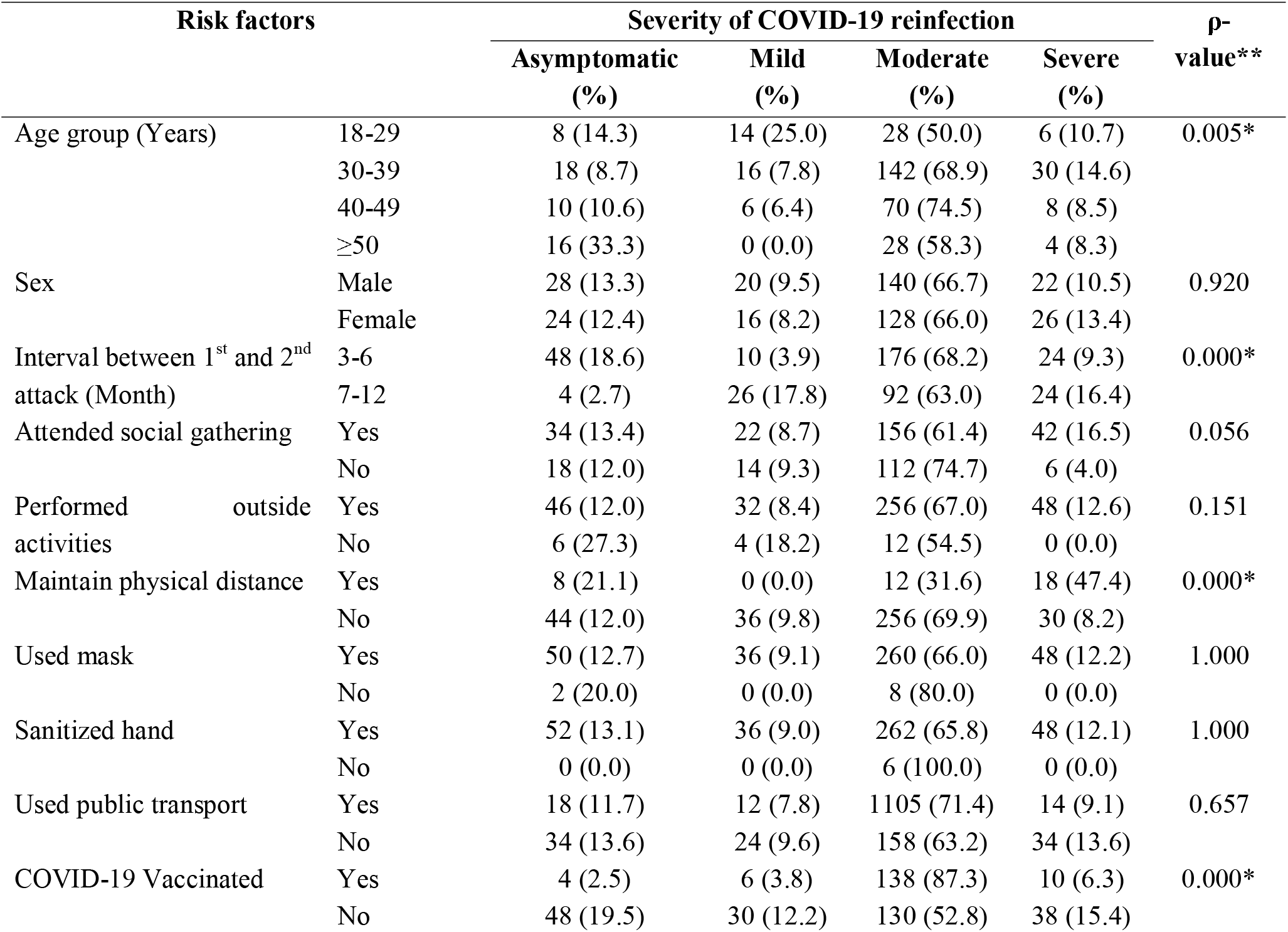
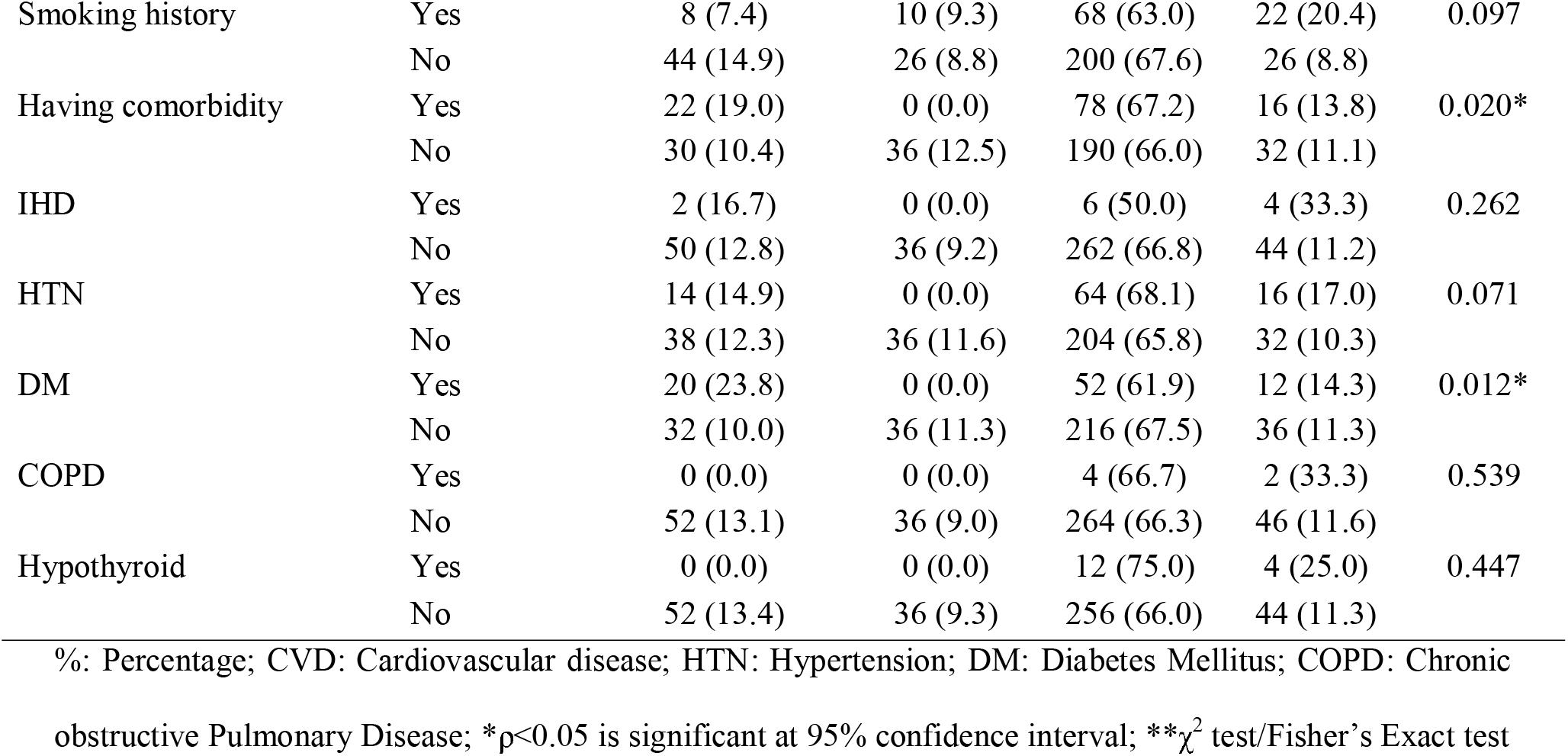
Association between risk factors and severity of COVID-19 reinfection.

| Risk factors | | Severity of COVID-19 reinfection | | | | ρ-value** |
| --- | --- | --- | --- | --- | --- | --- |
|  |  | Asymptomatic (%) | Mild (%) | Moderate (%) | Severe (%) |  |
| Age group (Years) | 18-29 | 8 (14.3) | 14 (25.0) | 28 (50.0) | 6 (10.7) | 0.005* |
|  | 30-39 | 18 (8.7) | 16 (7.8) | 142 (68.9) | 30 (14.6) |  |
|  | 40-49 | 10 (10.6) | 6 (6.4) | 70 (74.5) | 8 (8.5) |  |
| | $\geq 50$ | 16 (33.3) | 0 (0.0) | 28 (58.3) | 4 (8.3) | |
| Sex | Male | 28 (13.3) | 20 (9.5) | 140 (66.7) | 22 (10.5) | 0.920 |
|  | Female | 24 (12.4) | 16 (8.2) | 128 (66.0) | 26 (13.4) |  |
| Interval between 1<sup>st</sup> and 2<sup>nd</sup> attack (Month) | 3-6 | 48 (18.6) | 10 (3.9) | 176 (68.2) | 24 (9.3) | 0.000* |
|  | 7-12 | 4 (2.7) | 26 (17.8) | 92 (63.0) | 24 (16.4) |  |
| Attended social gathering | Yes | 34 (13.4) | 22 (8.7) | 156 (61.4) | 42 (16.5) | 0.056 |
|  | No | 18 (12.0) | 14 (9.3) | 112 (74.7) | 6 (4.0) |  |
| Performed outside activities | Yes | 46 (12.0) | 32 (8.4) | 256 (67.0) | 48 (12.6) | 0.151 |
|  | No | 6 (27.3) | 4 (18.2) | 12 (54.5) | 0 (0.0) |  |
| Maintain physical distance | Yes | 8 (21.1) | 0 (0.0) | 12 (31.6) | 18 (47.4) | 0.000* |
|  | No | 44 (12.0) | 36 (9.8) | 256 (69.9) | 30 (8.2) |  |
| Used mask | Yes | 50 (12.7) | 36 (9.1) | 260 (66.0) | 48 (12.2) | 1.000 |
|  | No | 2 (20.0) | 0 (0.0) | 8 (80.0) | 0 (0.0) |  |
| Sanitized hand | Yes | 52 (13.1) | 36 (9.0) | 262 (65.8) | 48 (12.1) | 1.000 |
|  | No | 0 (0.0) | 0 (0.0) | 6 (100.0) | 0 (0.0) |  |
| Used public transport | Yes | 18 (11.7) | 12 (7.8) | 1105 (71.4) | 14 (9.1) | 0.657 |
|  | No | 34 (13.6) | 24 (9.6) | 158 (63.2) | 34 (13.6) |  |
| COVID-19 Vaccinated | Yes | 4 (2.5) | 6 (3.8) | 138 (87.3) | 10 (6.3) | 0.000* |
|  | No | 48 (19.5) | 30 (12.2) | 130 (52.8) | 38 (15.4) |  |
| Smoking history | Yes | 8 (7.4) | 10 (9.3) | 68 (63.0) | 22 (20.4) | 0.097 |
|  | No | 44 (14.9) | 26 (8.8) | 200 (67.6) | 26 (8.8) |  |
| Having comorbidity | Yes | 22 (19.0) | 0 (0.0) | 78 (67.2) | 16 (13.8) | 0.020* |
|  | No | 30 (10.4) | 36 (12.5) | 190 (66.0) | 32 (11.1) |  |
| IHD | Yes | 2 (16.7) | 0 (0.0) | 6 (50.0) | 4 (33.3) | 0.262 |
|  | No | 50 (12.8) | 36 (9.2) | 262 (66.8) | 44 (11.2) |  |
| HTN | Yes | 14 (14.9) | 0 (0.0) | 64 (68.1) | 16 (17.0) | 0.071 |
|  | No | 38 (12.3) | 36 (11.6) | 204 (65.8) | 32 (10.3) |  |
| DM | Yes | 20 (23.8) | 0 (0.0) | 52 (61.9) | 12 (14.3) | 0.012* |
|  | No | 32 (10.0) | 36 (11.3) | 216 (67.5) | 36 (11.3) |  |
| COPD | Yes | 0 (0.0) | 0 (0.0) | 4 (66.7) | 2 (33.3) | 0.539 |
|  | No | 52 (13.1) | 36 (9.0) | 264 (66.3) | 46 (11.6) |  |
| Hypothyroid | Yes | 0 (0.0) | 0 (0.0) | 12 (75.0) | 4 (25.0) | 0.447 |
|  | No | 52 (13.4) | 36 (9.3) | 256 (66.0) | 44 (11.3) |  |

%: Percentage; CVD: Cardiovascular disease; HTN: Hypertension; DM: Diabetes Mellitus; COPD: Chronic obstructive Pulmonary Disease; \* $p < 0.05$ is significant at 95% confidence interval; \*\* $\chi^2$ test/Fisher's Exact test

**Table 5.** Multinomial Logistic regression of risk factors of severity of COVID-19 reinfection.

| Risk factors |  | Mild |  |  | Moderate |  |  | Severe |  |  |
| --- | --- | --- | --- | --- | --- | --- | --- | --- | --- | --- |
|  |  | B | AOR | ρ-value | B | AOR | ρ-value | B | AOR | ρ-value |
| Age group (Years) | 18-29 | - | - | - | -0.196 | 0.822 | 0.824 | 0.964 | 2.621 | 0.506 |
|  | 30-39 | - | - | - | 0.907 | 2.477 | 0.172 | 2.302 | 9.998 | **0.048** |
|  | 40-49 | - | - | - | 1.224 | 3.402 | 0.096 | 1.121 | 3.068 | 0.369 |
|  | ≥50 | Reference |  |  |  |  |  |  |  |  |
| Interval (Month) between 1<sup>st</sup> and 2<sup>nd</sup> attack | 3-6 | -3.488 | 0.031 | **0.000** | -2.025 | 0.132 | **0.017** | -2.827 | 0.059 | **0.002** |
|  | 7-12 | Reference |  |  |  |  |  |  |  |  |
| Maintained physical distance | Yes | - | - | - | -1.986 | 0.137 | **0.013** | 0.938 | 2.556 | 0.232 |
|  | No | Reference |  |  |  |  |  |  |  |  |
| COVID-19 Vaccinated | Yes | 0.898 | 2.455 | 0.405 | 2.781 | 16.127 | **0.001** | 1.359 | 3.894 | **0.047** |
|  | No | Reference |  |  |  |  |  |  |  |  |
| Having comorbidity | Yes | - | - | - | 0.140 | 1.151 | 0.907 | 0.203 | 1.225 | 0.896 |
|  | No | Reference |  |  |  |  |  |  |  |  |
| Diabetes Mellitus | Yes | -0.127 | 0.881 | 1.000 | -0.909 | 0.403 | 0.446 | -0.396 | 0.673 | 0.797 |
|  | No | Reference |  |  |  |  |  |  |  |  |

Reference category: Asymptomatic; AOR= Adjusted Odds Ratio; CI: Confidence interval; \*p<0.05, significant at 95% CI; B=Regression coefficient

The severity of COVID-19 infection was significantly associated with comorbidity (ρ = 0.020). The study depicted that 67.2% and 13.8% of the patients having comorbidity had moderate and severe infections respectively. The study showed that the severe infection was significantly (ρ = 0.012) higher in the patients with DM (14.3%) than in the patients without DM (11.3%). The severity of reinfection was also significantly associated with the COVID-19 vaccination status of the patients (ρ<0.001). The study found that the severe infection was significantly higher in the unvaccinated (15.4%) than in the vaccinated (6.3%) patients. On the other hand, the moderate infection was significantly higher in the vaccinated (87.3%) than in the unvaccinated (52.8%) patients (Table-1).

Multi-nominal logistics regression analysis revealed that three to six months’ interval between first and second incidents had less chance of having mild (AOR=0.031, ρ=0.000), moderate (AOR=0.132, ρ=0.017) and severe (AOR=0.059, ρ=0.002) infection. The reinfected patients who maintained physical distance had less chance of having moderate severity of infection (AOR=0.137, ρ=0.013). The study revealed that the patients who were vaccinated had higher chance of having moderate (AOR=16.127, ρ=0.001) and severe (AOR=3.894, ρ=0.047) infection (Table-1).

## Discussion

Reinfections with SARS-CoV-2 have been evident as an increasing concern during the COVID-19 pandemic era. This sizzling public health problem is not well-addressed. This inventive countrywide cross-sectional study aimed to identify the proportion of COVID-19 reinfection along with the severity and associated risk factors. The study explored selected clinical attributes, background features, exposure, preventive practices, comorbidities, and vaccination status of the victims. This current initiative endures a potential policy importance to devise preventive strategies for COVID-19 reinfection.

Based on the data collected from HIU of DGHS, Bangladesh, the estimated proportion of COVID-19 reinfection was 1.14%. In this regard, the study conducted in France during the period from June 2020 to January 2021, reported a little lower (0.47%) rate of reinfection [2]. This variation could be justified by the difference in study context and study participants.

In the present study, the mean (±SD) age of reinfected patients was 37 (±8.5) years. A little difference was found in the rate of reinfection between male and female patients (male=52.0%; female=48.0%). The study conducted in France showed that mean (±SD) age was 50 (±22) years, and 51.2% were male [2]. The dissimilarity of the age of reinfected patients was evident between the two studies, which could be due to the variation of immunity status of the people of different ages in the two countries. Both the studies showed that an almost equal percentage of male and female participants were reinfected, which indicates that gender differences didn’t make any influence on the occurrence of the reinfection. The present study revealed that the majority of the reinfected patients (37.1%) were healthcare workers. It could be argued that the healthcare workers were employed in diverse health facilities, where they had to deal with COVID-19 patients and became exposed to sources of infection, which increased their vulnerability for reinfection. The study found that almost all (98.0%) of the patients were hailing from urban settings. In this respect, another study conducted in Bangladesh also found a higher COVID-19 infection rate in urban areas [15]. These findings indicate that both infection and reinfection rates were higher in urban areas. It could be claimed that the probability of disease transmission was higher in the urban areas due to high population density, overcrowded workplaces, industrialization, public transport use, and diverse religious and social gatherings. This study finding invites comprehensive preventive interventions for the urban-dwelling COVID-19 patients to prevent them from reinfection.

The current study portrayed that 88.1% of the patients had oxygen saturation ≥90%, and the majority (90.6%) were treated at their homes. It could be explained by the fact that the majority of the reinfected patients had mild to moderate degrees of infection, which compelled them to take hospitalized treatment. It was unveiled that 63.9% of the patients had an interval of 3-6 months between first and second incidents of COVID-19 infections. The study conducted in France identified 3-10 months elapsed between the first and the second episodes of COVID-19 infections [2]. The results of both the studies indicate that possibility of reinfection was more during 3-6 months since the first attack of COVID-19. In respect of severity of reinfection, the present study exposed that the probability of mild, moderate, and severe infection was significantly lower than asymptomatic infection among patients who had reinfection within 3-6 months in comparison to 7-12 months after the first incident. This study finding indicates that the severity of reinfection increased with increasing intervals as immunity to reinfection decreased with increasing intervals between first and second episodes of infections. This finding recommends strict and sincere preventive measures to the COVID-19 patients for abiding during three to six months since the first incident to prevent the reinfection and its severity.

The study depicted that the majority (94.6%) of the reinfected patients performed outside activities, 62.9% attended the social gathering, 38.1% used public transport, and 90.6% didn’t maintain physical distance after recovery from the first incident of COVID-19 infection. It could be justified by the fact that the people assumed that there was no need for protective measures after the first infection. Some of the studies [16,17] also proposed that COVID-19 infection itself provides naturally acquired immunity to the patient and vaccination boost up immunity. But our study revealed that poor maintenance of physical distance was significantly associated with the severity of COVID-19 reinfection, and the patients who maintained physical distance had less chance of having a moderate infection. This finding forwards a strong recommendation for maintaining social distance even by the previously infected COVID-19 patients to avert reinfection.

The current study also revealed that 28.7% patients had one or more comorbidities including hypertension (23.3%), diabetes mellitus (20.8%), and chronic obstructive pulmonary disease (1.5%). Another study conducted in Wuhan, China also determined those comorbidities in different proportion and found that 40% patients had hypertension, 20% had diabetes mellitus and 13% had chronic obstructive pulmonary disease [18]. These differences could be due to the disparity of social, environmental, cultural, and lifestyle between the two countries. But both the studies didn’t reveal any associating of the severity of COVID-19 reinfection with comorbidities.

Our study found the majority (60.9%) of the reinfected patients were unvaccinated against COVID-19. A significant association was evident between the severity of reinfection and the vaccination status of the COVID-19 patients. The present study noticed that the vaccinated patients had significantly more chance of having moderate and severe infection than the unvaccinated patients. But the study conducted in Kentucky acclaimed that vaccination can reduce the risk of COVID-19 reinfection [19]. Our extensive literature confronted an enormous scarcity of relevant studies concerning the association between vaccination status and severity of COVID-19 reinfection. Though vaccination may reduce the risk of COVID-19 reinfection but our study indicates that it may increase the possibility of flaring up symptoms in reinfected cases. So, the current study applauds further analytical studies to assess the relationship between the severity of reinfection and the vaccination status of the patients.

Despite the limitation of telephone interview, our study findings provide new insight on the proportion of COVID-19 reinfection along with its severity and associated risk factors. The study also portrays information on the association of the severity of reinfection with preventive practices and the vaccination status of patients. The study findings could contribute to designing an efficient preventive algorithm to alleviate COVID-19 reinfection in a more pragmatic approach. The study conserves decisive policy implications for devising effective interventions to prevent the severity of COVID-19 reinfection. Moreover, the study inspires future inclusive studies on the COVID-19 reinfection and offers a scope of comparison considering geographical, demographical, socio-cultural, epidemiological, and clinical determinants.

## Conclusion

Our findings indicate that the COVID-19 reinfection is associated with poor social distancing and the vaccination status of the patients. To prevent reinfection, patients must abide by preventive measures even after recovery and vaccination. However, future research efforts are required to carry out analytical studies to correlate vaccination with the severity of COVID-19 reinfection.

## Data Availability

The datasets generated and/or analyzed during the current study are not publicly available as the ethical vote did not include open data access but are available from the corresponding author on reasonable request.

## Authors’ contributions

MZI: Contributed to study conception, study design, data acquisition, interpretation of data, preparation, and reviewing the manuscript. MZI had full access to all of the data in the study and takes responsibility for the integrity of the data and the accuracy of the data analysis. BKR: Contributed to study design, interpretation of data, preparing, and reviewing the manuscript. SAA: Contributed to study design, data acquisition, data analysis, interpreting results, preparation, and reviewing the manuscript. SF: Contributed to study design, data acquisition, data analysis, interpretation of results, and reviewing the manuscript. SSE: Contributed to data acquisition, data analysis, interpretation of results, and reviewing the manuscript. MAK: Contributed to data acquisition, interpretation of results, and reviewing the manuscript. Each author has approved the submitted version of the manuscript and has agreed to be personally accountable for the author’s own contributions and to ensure that questions related to the accuracy or integrity of any part of the work, even ones in which the authors were not personally involved, are appropriately investigated, resolved and the resolution documented in the literature.

## Consent to publish

The manuscript does not contain any person’s data in any form including individual details, images, or videos. So, we didn’t obtain consent for publication from the patients.

## Ethical approval

Ethical approval of the study was given by the Institutional Review Board of the National Institute of the Preventive and Social Medicine (NIPSOM), Dhaka, Bangladesh with reference no. NIPSOM/IRB/2021/06, date: 02/03/2021. Keeping compliance with Helsinki Declaration for medical research involving human subjects, we obtained informed consent (written or verbal) from every participant by explaining the study design, purpose, procedure, risk, and benefits before the interview. We maintained privacy, anonymity, and confidentiality of data strictly.

## Availability of data and material

Availability was only given to the patients for their information and specific members of the study. The datasets generated and/or analyzed during the current study are not publicly available as the ethical vote did not include open data access but are available from the corresponding author on reasonable request.

## Acknowledgements

All authors would like to acknowledge the staff of the Health Information Unit of DGHS, Bangladesh for their tenacious and unrestricted support in sharing patients’ records. We would like to forward sincere appreciation to the COVID-19 patients and their families for their unconditional assistance in data collection through a telephone interview.

